# Efficacy and Safety of Prehospital Tranexamic Acid for Trauma: A Systematic Review and Meta-Analysis

**DOI:** 10.1101/2025.03.15.25324013

**Authors:** Mario Bolarte-Arteaga, Julian Espinoza-Portilla, Flor Santa Cruz-De Lama, Carlos Zavaleta-Corvera

## Abstract

**Background:** Trauma is one of the leading causes of death worldwide. Tranexamic acid (TXA) has shown effectiveness in reducing hemorrhage-related mortality in a hospital setting. However, its application in the prehospital setting still presents challenges.

**Aim:** To determine the efficacy and safety of TXA administered in the prehospital setting in trauma patients.

**Methods:** A systematic review with meta-analysis was conducted. PRISMA guidelines were followed.Randomized clinical trials evaluating the administration of TXA in the prehospital setting in trauma patients aged 18 years or older were included. Studies were assessed by two reviewers independently. The GRADE approach was used to assess the quality of evidence and ROB2 to identify the risk of bias. Results were analyzed by meta-analysis, using fixed or random effects models, depending on the heterogeneity observed.

**Results:** A total of 979 records were identified; PubMed (149), Embase (565), Cochrane (29) and Scopus (236). Three studies were included. After analysis TXA reduced mortality in the first 24 hours (RR 0.74, 95% CI: 0.56-0.97; P = 0.03) and at 28 days (RR 0.82, 95% CI: 0.69-0.98; P = 0.03). No improvement in survival with favorable long-term functional outcome was observed (RR 1.11, 95% CI: 0.91-1.35; P = 0.29). No significant differences were found in adverse events such as deep vein thrombosis (RR 1.23, 95% CI: 0.96-1.58; P = 0.11), pulmonary embolism (RR 1.08, 95% CI: 0.76-1.53; P = 0.66) or myocardial infarction (RR 2.17, 95% CI: 0.75-6.31; P = 0.15).

**Conclusions:** Prehospital TXA use in trauma patients reduces short-term mortality, mortality in the first 24 hours, and mortality at 28 days. In addition, it does not increase the risk of serious adverse events; deep vein thrombosis, pulmonary embolism, myocardial infarction, or ischemic stroke.

## INTRODUCTION

Severe trauma is one of the leading causes of death worldwide. Young people are one of the most affected, accounting for 10% of the global burden of disease (1). According to data from the World Health Organization (WHO), injuries caused by traffic accidents, falls and injuries resulting from secondary trauma or violent episodes are responsible for more than 5 million deaths each year. In percentage terms, this figure represents almost 9% (2). A large percentage of these deaths occur in low- and middle-income countries, where prehospital care systems are often less developed (3).

Hemorrhage control is critical in the management of patients with severe trauma, as uncontrolled bleeding is the leading cause of preventable death (4). Tranexamic acid (TXA) has been identified as a key tool in this setting due to its ability to reduce excessive fibrinolysis, a molecular process that contributes to massive blood loss following severe trauma (5). At the molecular level, TXA acts by inhibiting the binding of plasminogen to fibrin, thereby preventing the formation of plasmin, an enzyme responsible for the breakdown of blood clots (6).

The mechanism of action of TXA is crucial in stabilizing clots formed in response to injury, thereby aiding active bleeding (7). This antifibrinolytic effect is particularly beneficial in the prehospital setting, where the time to definitive surgical intervention may be prolonged, and early control of bleeding may make the difference between life and death (8). Administration of TXA in the prehospital setting has been supported by studies such as the CRASH-2 trial, which showed a significant reduction in all-cause mortality in patients receiving TXA within the first three hours after trauma (9).

Furthermore, the epidemiological bases indicate that severe trauma is not only a public health problem, but also a significant economic burden, with direct and indirect costs amounting to billions of dollars annually worldwide (10). Implementation of cost-effective treatments such as TXA in the prehospital phase could not only save lives, but also reduce costs associated with long-term emergency care (11).

The application of TXA in the prehospital setting faces challenges. These include the lack of standardized protocols, variability in its use among different emergency systems, and the lack of adequate logistics as well as human resources for prompt care (12). Although evidence supports its safety, studies suggest the need to further evaluate its potential risk of associated thromboembolic events, especially in high-risk populations (13).

Continued research is essential to optimize the use of TXA, including determining the most effective doses, the most appropriate timing of administration, and its impact on different patient subgroups (14,15).

Acharya P, et al. (UK, 2023) conducted a systematic review to determine the efficacy of using TXA in the treatment of trauma patients. However, not all relevant clinical outcomes such as long-term function or other potentially important adverse effects were included. Although their focus was primarily on short-term mortality, they left out outcomes that could be critical to comprehensively assess the safety and efficacy of TXA in a broader context, thus limiting the interpretation of clinical results (16). Thus, the aim of the present study is to determine the efficacy on hemodynamic outcomes in the prehospital setting of trauma patients.

## METHODS

### Study design

This study followed a systematic review design with meta-analysis, adhering to the PRISMA (Prefered Reporting Items for Systematic Review and Meta-Analyses) guidelines (20). The following protocol was registered in PROSPERO.

### Research question

The objective of this review was to assess the efficacy and safety of tranexamic acid (TXA) administration in the prehospital management of patients at significant risk of bleeding following trauma.Patients aged 18 years or older who have experienced trauma with significant risk of bleeding and have received prehospital care were included. Studies that included the administration of TXA in the prehospital setting compared to standard prehospital care were evaluated. All available outcomes in the selected research articles related to efficacy and safety were evaluated.

### Search strategy

A comprehensive search was conducted in electronic databases such as Pubmed, Embase Cochrane library and Scopus. No language or publication date restrictions were applied. Search terms will include “Prehospital”, “Emergency Medical Services”, “Out-of-Hospital Care”, “Prehospital Care”, “Tranexamic Acid”, “TXA”, “Antifibrinolytic patients”, “trauma”, “Injury”, “Trauma Hemmorrhage” (Appendix 1). In addition, Boolean operators such as AND and OR were added. Reference lists of included studies were reviewed to identify additional studies with potential to be entered into the analysis until August 31, 2024.

### Inclusion criteria

Studies were included as randomized controlled trials (RCTs) that evaluated the administration of TXA in the prehospital setting or before arrival at the hospital in patients older than 18 years who have suffered trauma. Comparators were those who had received any type of prehospital care without the use of TXA.

### Exclusion criteria

Observational studies such as case-control, cohort or cross-sectional studies, as well as descriptive studies, case reports, case series, editorials or articles that do not have complete information such as abstracts. Secondary studies such as narrative reviews, systematic reviews with or without meta-analysis, among others, were also excluded. Articles that do not provide comparative data between patients who received TXA. Studies that have used the intervention outside the prehospital context.

### Selection of studies

Titles and abstracts of articles identified by the initial search were assessed by two independent reviewers. Potentially relevant studies were reviewed in full text. Discrepancies were resolved by discussion or, if necessary, by consulting a third expert reviewer.

### Data Extraction

Data extraction was performed by two reviewers independently, using a standardized data extraction table that will include the following variables: demographic data; age, sex, type of trauma, details of the intervention; dose of TXA administered, time until administration from the traumatic event, as well as a description of the primary outcomes and those considered in each study as appropriate.

### Bias risk assessment and quality

The quality of the included studies was assessed using the Cochrane tool for risk of bias in randomized trials (Rob 2.0). The following domains were considered; Allocation, blinding, incomplete outcome data and other sources of bias.

### Evaluating the quality of evidence

The quality of evidence for each of the outcomes studied was assessed using the GRADE (Grading of Recommendations, Assessment, Development, and Evaluation) approach. GRADE classifies the quality of evidence into 4 levels: High, moderate, low, and very low, based on factors such as risk of publication bias. This approach helped to interpret the strength of recommendations derived from the review findings.

### Statistical analysis

A meta-analysis was performed to calculate the risk ratio (RR) with 95% confidence intervals (CI) for binary outcomes. A random-effects model was used if significant heterogeneity of the outcomes was observed.(I^2^ > 50%);Otherwise, a fixed-effect model was used. Heterogeneity between studies was assessed using the I2 statistic and the Chi2 test. Sensitivity and subgroup analysis were performed to explore sources of heterogeneity.

### Qualitative synthesis

In addition to the meta-analysis, a qualitative synthesis was performed for those studies or outcomes where a meta-analysis is not possible due to heterogeneity of the studies or lack of comparable quantitative data.

### Ethical considerations

This study was a secondary study. Therefore, it does not require the approval of an ethics committee for the development of this research.

## RESULTS

A total of 979 records were identified through searching databases and manual records. Databases such asPubMed (149), Embase (565), Cochrane (29) and Scopus (236). Subsequently, 433 duplicate records were eliminated and 537 were excluded for not meeting the pre-established criteria for the review. 9 articles were selected for full text review, resulting in the exclusion of 6 studies; 5 of them for being outside the type of study specified in the criteria and one because it did not correspond to the population of interest (Figure 1).

**Figure 1.**
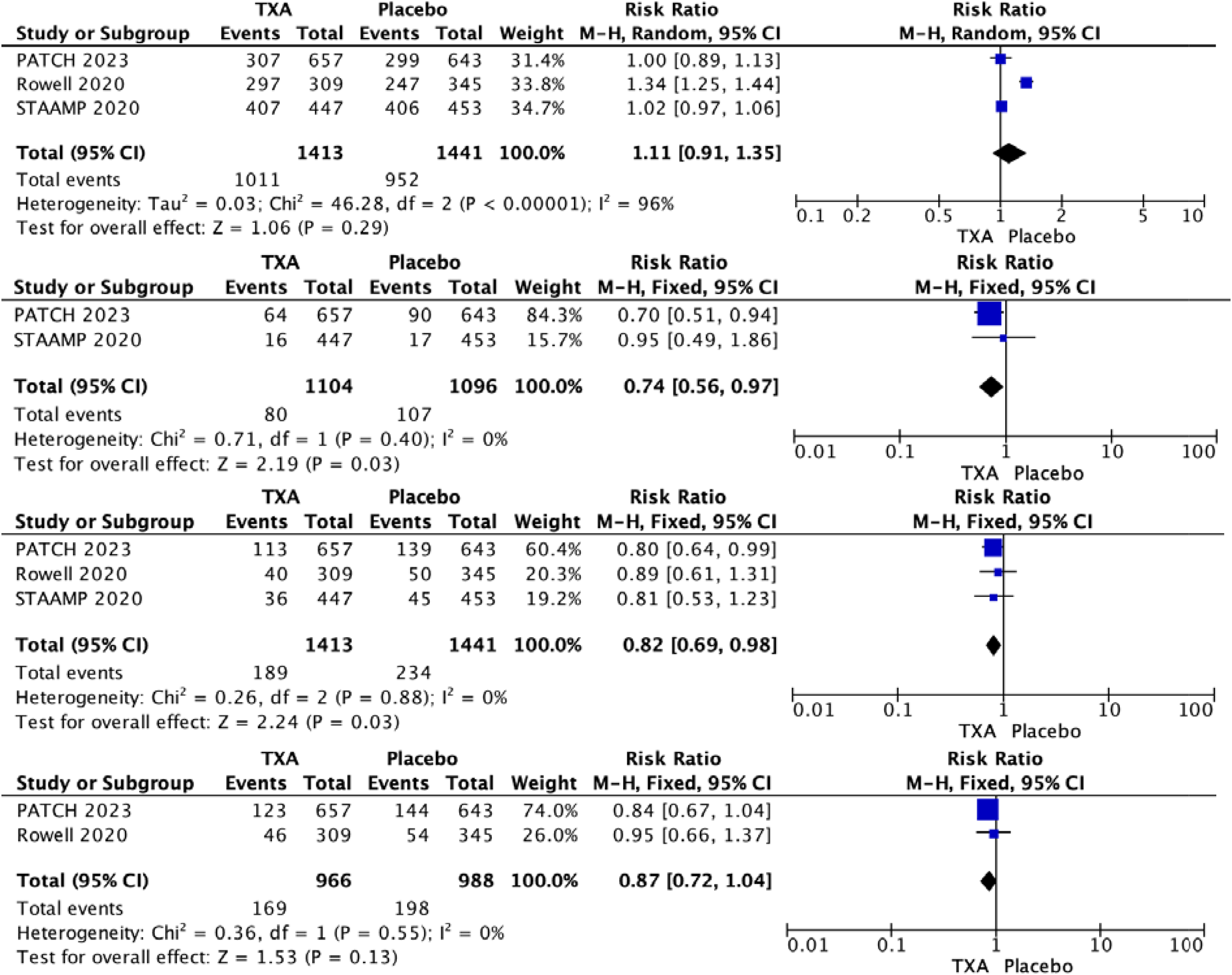
**Efficacy outcomes related to TXA and prehospital administration for trauma. A. Survival with a favorable functional outcome B. Death 24 hours after injury C. Death 28 days after injury D. Death 6 months after injury**

Thus, 3 studies were included in the systematic review and meta-analysis. All of them were randomized controlled clinical trials (RCTs) that analyzed the use of tranexamic acid (TXA) in patients with different types of trauma (moderate, severe). In the PATCH study (2023), patients in the study received 1 g of TXA before hospital admission, followed by an infusion of 1 g for 8 hours. In the STAAMP study (2020), the intervention consisted of an infusion of 1 g of TXA for 8 hours or a bolus followed by an infusion of 1 g of TXA. In Rowell et al. (2020), the intervention was based on the administration of a bolus of 1 g of TXA followed by an infusion or a bolus of 2 g of TXA without additional infusion (Table 1).

**Table 1.** Baseline Characteristics of Included Studies.

| Study | Country | Type of study | Sample size | Age (mean ± SD) | Male, n (%) | Injury Type (Blunt / Penetrating) | Out-of-hospital Glasgow Coma Scale score, mean (SD) | TXA Doses | BP (mmHg) | Heart Rate (bpm) | Time to TXA/Placebo Administration |
| --- | --- | --- | --- | --- | --- | --- | --- | --- | --- | --- | --- |
| **PATCH, 2023 (17)** | Australia | RCT | 1300 | 44.1 ± 19.7 / 44.2 ± 18.9 | 459 (69.9) / 459 (71.4) | Blunt: 92.8% / 91.4%,<br>Penetrating: 6.7% / 8.6% | <9: 229/655 (35.0) 211/642 (32.9), 9 - 12: 51/665 (7.8) 73/642 (11.4), 13 - 15: 375/655 (57.3) 358/642 (55.8) | 1 g before hospital admission, followed by a 1-g infusion over a period of 8 hours after arrival at the hospital | ≤75 mmHg: 38.5% / 39.6%,<br>76–89 mmHg: 34.1% / 31.5%,<br>≥90 mmHg: 27.4% / 28.9% | NEITHER | <1 hr: 32.6% / 27.5%, 1-2 hrs: 45.3% / 51.3%,<br>≥2 hrs: 22.1% / 21.2% |
| **STAAMP, 2020 (18)** | USA | RCT | 900 | 42 ± 18 / 42 ± 18 | 330 (74.0) / 356 (78.1) | Blunt: 61.5% / 63.3%,<br>Penetrating: 36.5% / 36.2% | Initial Glasgow Coma Scale score <8: 107 (23.5) 89 (19.9) | Intervention arm patients received no additional tranexamic acid, 1 g of tranexamic acid infused for 8 hours, or a bolus of 1 g of tranexamic acid followed by 1 g of tranexamic acid infused for 8 hours. | SBP ≤ 70 mmHg: 18.5% / 35.5% | 117.7 ± 21.5 / 119.4 ± 20.5 | <1 hr: 30.6% / 28.2%, 1-2 hrs: 49.4% / 48.6%,<br>≥2 hrs: 20.0% / 23.2% |
| **Rowell, 2020 (19)** | USA | RCT | 966 | 40 (26-56) / 36 (25-55) | 255 (74) / 233 (75) | Blunt 98% / 95%,<br>Penetrating: 1% / 5% | 7.8 (3.3) / 7.6 (3.2) | Bolus maintenance group: 1-g IV tranexamic acid bolus in the out-of-hospital setting, followed by a 1-g tranexamic acid IV infusion initiated upon hospital arrival and infused over 8 hours. Bolus only group: 2-g IV tranexamic acid bolus in the out-of-hospital setting, followed by a placebo infusion. Placebo group: IV placebo bolus in the out-of-hospital setting, followed by an IV placebo infusion. | NEITHER | NEITHER | 40 to 43 minutes across groups: bolus completion percentage ranged from 93% to 95%. |

**Abbreviations:** RCT: Randomized Controlled Trial; SD: Standard Deviation; n: Number; TXA: Tranexamic Acid; BP: Blood Pressure; bpm: Beats Per Minute; NI: Not Indicated; mmHg: Millimeters of Mercury; SBP: Systolic Blood Pressure; IV: Intravenous.

**Table 4.** GRADE evidence for Quality of the studies included in the review.

| No. of studies | Study design | Risk of bias | Inconsistency | Indirectness | Vagueness | Other considerations | Drug X | Standard Care | Relative (95% CI) | Absolute (95% CI) | Quality | Importance |
| --- | --- | --- | --- | --- | --- | --- | --- | --- | --- | --- | --- | --- |
| 3 | RCT | Low | None | None | Minimal | None | PATCH 2023 | Placebo | 1.11 (0.91-1.35) | Not significant | ●●●● | Critical |
| 2 | RCT | Low | Minimal | None | Some | None | STAAMP 2020 | Placebo | 1.09 (0.75-1.59) | Not significant | ●●●○ | Important |
| 2 | RCT | Low | Significant | None | Significant | None | Rowell 2020 | Placebo | 1.22 (0.68-2.20) | Not significant | ●●●○ | Important |

Legend: ●●●●= High Quality, ●●●○= Moderate Quality, ●●○○= Low Quality, ●○○○= Very Low Quality.

### Survival with a favorable functional outcome

Survival and/or functional outcomes were evaluated in patients who received prehospital TXA. TXA was found to bedid not result in a significant improvement in survival with a favorable functional outcome compared with placebo, with a RR of 1.11 (95% CI: 0.91, 1.35; P = 0.29). Although the Rowell 2020 study reported a significant improvement (RR 1.34, 95% CI: 1.25, 1.44), the PATCH 2023 and STAAMP 2020 studies found no relevant differences. The high heterogeneity (I^2^ = 96%) suggests considerable variability between studies, affecting the interpretation of the variable (Figure 1A).

### Death 24 hours after injury

Mortality was assessed within 24 hours after trauma.The analysis showed that TXA administration significantly reduced the risk of death within the first 24 hours, RR 0.74 (95% CI: 0.56, 0.97; P = 0.03). Heterogeneity was low (I^2^ = 0%), suggesting consistency across the included studies (Figure 1B).

### Death 28 days after injury

Analysis of mortality at 28 days after trauma showed that TXA was associated with a significant reduction in the risk of death compared with placebo. The data showed a RR of 0.82 (95% CI: 0.69, 0.98; P = 0.03). Heterogeneity was low (I^2^ = 0%), suggesting that the results were consistent across the included studies (Figure 1C).

### Death 6 months after injury

Analysis of mortality at 6 months after trauma did not show a significant reduction in the risk of death with the use of TXA, with a RR of 0.87 (95% CI: 0.72, 1.04; P = 0.13). In the PATCH 2023 study, the RR was 0.84 (95% CI: 0.67, 1.04), while in the Rowell 2020 study it was 0.95 (95% CI: 0.66, 1.37). Heterogeneity was low (I^2^ = 0%), indicating consistency across studies. However, the overall effect was not statistically significant (P = 0.13), suggesting that TXA has no clear impact on reducing long-term mortality in trauma patients (Figure 1D).

### Deep venous thrombosis

The incidence of deep vein thrombosis (DVT) was assessed in patients who received prehospital TXA. TXA was found to result in no significant difference in the incidence of DVT compared with placebo, with a pooled RR of 1.23 (95% CI: 0.96, 1.58; P = 0.11). Although the STAAMP 2020 study reported an elevated risk (RR 1.74, 95% CI: 0.69, 4.37), the PATCH 2023 and Rowell 2020 studies found no relevant differences. Low heterogeneity (I^2^ = 0%) suggests low variability between studies, strengthening the reliability of the results for this variable (Figure 2A).

**Figure 2.**
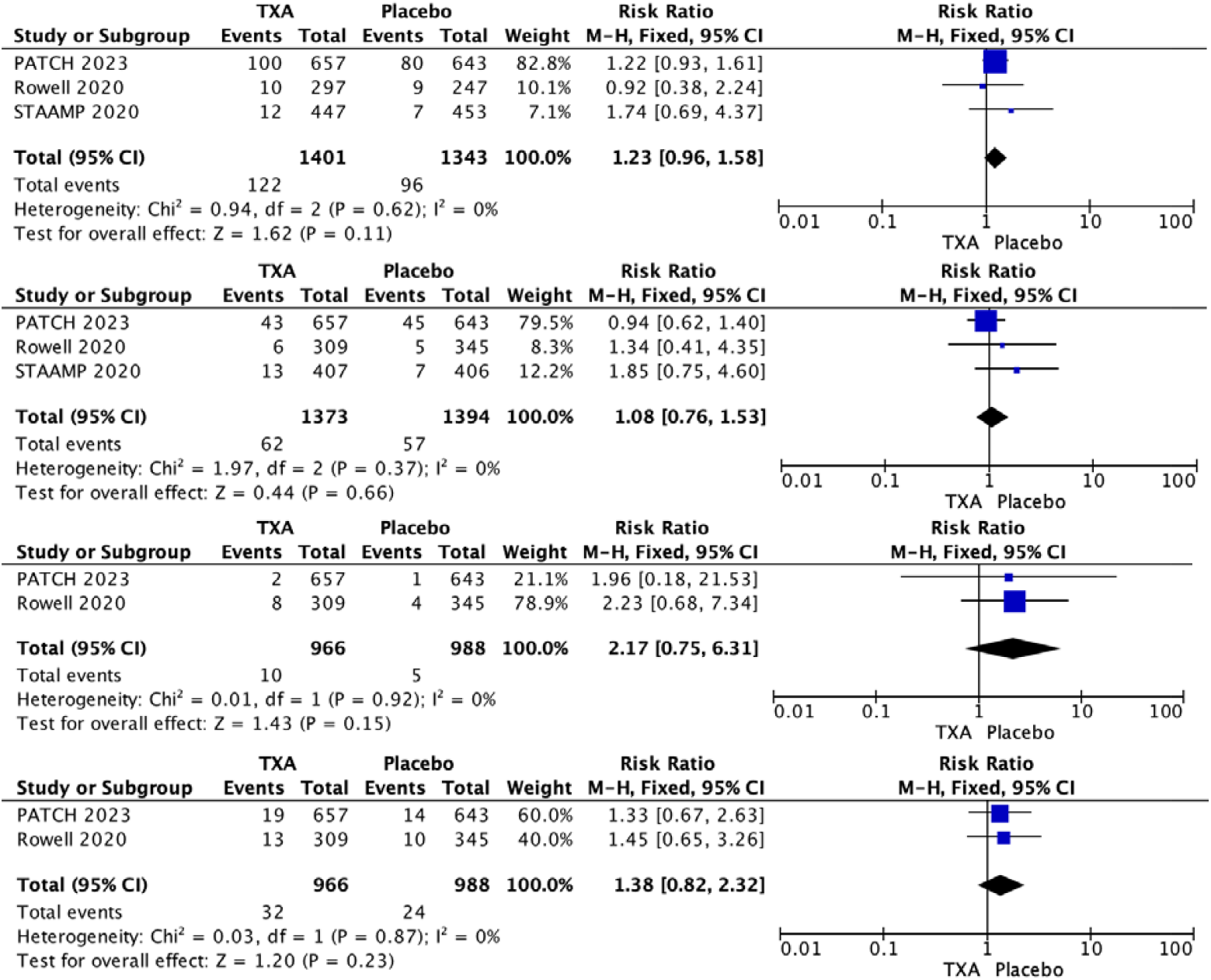
**Safety outcomes related to TXA and prehospital administration in trauma A. Deep venous thrombosis B. Pulmonary embolism C. Myocardial infarction D. Ischemic Stroke**.

### Pulmonary embolism

The incidence of pulmonary embolism (PE) was assessed in patients who received prehospital TXA. TXA was found to result in no significant difference in the incidence of PE compared with placebo, with a pooled RR of 1.08 (95% CI: 0.76, 1.53; P = 0.66). Although the STAAMP 2020 study showed an increased risk of pulmonary embolism (RR 1.85, 95% CI: 0.75, 4.60), the PATCH 2023 and Rowell 2020 studies found no relevant differences. Low heterogeneity (I^2^ = 0%) suggests little variability between studies, strengthening the reliability of the results for this variable (Figure 2B).

### Myocardial infarction

The incidence of myocardial infarction was assessed in patients given prehospital TXA. TXA was found to result in no significant difference in the incidence of myocardial infarction compared with placebo, with a pooled RR of 2.17 (95% CI: 0.75, 6.31; P = 0.15). The PATCH 2023 and Rowell 2020 studies reported an increased risk of myocardial infarction with TXA use, but without statistical significance (PATCH 2023: RR 1.96, 95% CI: 0.18, 21.53; Rowell 2020: RR 2.23, 95% CI: 0.68, 7.34). Low heterogeneity (I^2^ = 0%) indicates consistency between studies, which reinforces the reliability of the results for this variable (Figure 2C).

### Ischemic Stroke

The incidence of ischemic stroke was assessed in patients given prehospital TXA. TXA was found to result in no significant difference in the incidence of ischemic stroke compared with placebo, with a pooled RR of 1.38 (95% CI: 0.82, 2.32; P = 0.23). The PATCH 2023 and Rowell 2020 studies both showed an increased risk, although without statistical significance (PATCH 2023: RR 1.33, 95% CI: 0.67, 2.63; Rowell 2020: RR 1.45, 95% CI: 0.65, 3.26). Low heterogeneity (I^2^ = 0%) suggests good consistency across studies, supporting the reliability of the results for this endpoint (Figure 2D).

### Risk of Bias ROB2.0 tool

Risk of bias was assessed using the RoB2 tool. Low risk of bias was observed in most domains assessed, including random sequence generation and allocation concealment. In addition, the studies showed a low risk of performance bias, except for one area in the STAAMP 2020 study, where there was some uncertainty in outcome assessment, leading to an unclear risk of bias in outcome assessment. In terms of attrition bias due to incomplete data, all three studies were at low risk (Figure 3).

**Figure 3.**
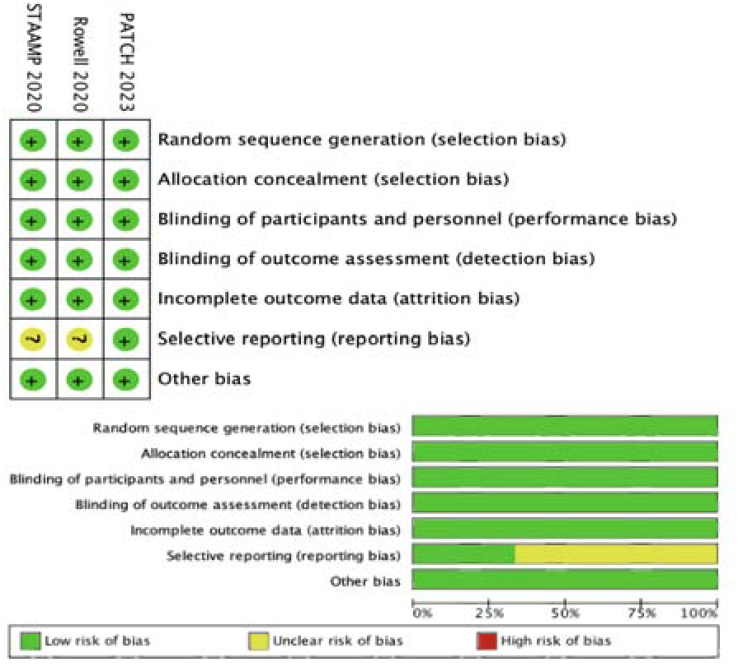
Quality assessment of included studies ROB2.0 (draft)

### GRADE evidence

The results of the GRADE analysis of the three included studies demonstrate variable quality of evidence (17,18,19). The PATCH 2023 study was assessed as high quality, indicating that the results are consistent, accurate, and directly applicable to the research question, with no significant risk of bias or imprecision. The STAAMP 2020 and Rowell 2020 studies showed moderate quality, due to slight inconsistency and higher imprecision in the results (Figure 4).

## DISCUSSION

After analysis of the available data, the results of the included studies were consistent. The results showed low heterogeneity overall. These findings suggest robustness regarding the efficacy and safety of TXA. TXA may apparently be beneficial in the acute phase of trauma management, however, its long-term impact and survival requires further investigation. Tentatively, because multiple clinical and sociodemographic factors may be related to long-term mortality, these may negatively influence mortality in trauma patients.

Our meta-analysis reveals a significant reduction in 24-hour and 28-day mortality with the use of TXA but in the long term, it would not have a statistically significant impact on 6-month mortality. El-Menyar et al. (Qatar, 2018), reported a relative risk (RR) of 0.82 (95% CI: 0.70-0.96) in the reduction of 28-day mortality, with no clear impact on thromboembolic events (21).)Similarly, Biffi et al. (Italy, 2023) observed a RR of 0.79 (95% CI: 0.66-0.94) in short-term mortality in patients with significant bleeding, but found no difference in thromboembolic events.(22)Stansfield et al. (UK, 2020) also reported a reduction in mortality with TXA, although with inconsistent results regarding thromboembolic events.(23).

Thus, while it is true that TXA can produce a decrease in short-term mortality without efficacy on long-term mortality, it is important to mention that there are factors independent of TXA consumption that can darken the prognosis of trauma patients and that were not considered in the analysis of the evaluated studies. Haider et al. (USA, 2020), evaluated the long-term results of 1,736 patients with aInjury Severity Score (ISS) ≥9, six to twelve months after experiencing moderate to severe trauma. 62% of patients reported current physical limitations, while 37% needed assistance for at least one daily activity. 20% showed symptoms of post-traumatic stress disorder (PTSD). In addition, 41% of patients who were working before the trauma were unable to return to work (p < 0.05) (24). González-Robledo et al. (Spain, 2015) conducted a retrospective and longitudinal study with the aim of identifying the prognostic factors associated with mortality in patients with severe trauma, age over 65 years (OR 3.15), cranial injuries (OR 3.1), pupillary abnormalities (OR 113.88), a score on the Glasgow coma scale less than or equal to 8 (OR 12.97), and serum lactate levels greater than 4 mmol/L (OR 9.7) were those associated with an increase in mortality (25).

The evaluation of TXA in the prehospital environment is essential because different clinical contexts can also affect negative outcomes related to safety. Thus, in the present study, no association was found between TXA consumption and deep vein thrombosis, pulmonary embolism, myocardial infarction or ischemic cerebrovascular events.Zhang and Liu (2024) evaluated the safety in patients with head trauma. The results showed that the use of TXA did not increase the risk of significant adverse events, including vascular occlusive events (RR 0.85, p = 0.16) and pulmonary embolism (RR 0.76, p = 0.26). In addition, no increase in seizures (RR 1.11, p = 0.27) or hemorrhagic complications (RR 0.78, p = 0.14) was observed (26). However, Mahdi Al-Jeabory et al. (Poland, 2021) performed a systematic review and meta-analysis to evaluate the safety of TXA in emergency trauma. The risk of a vascular occlusive event in the TXA-treated group was found to be 1.8%, compared to 2.1% in the non-TXA-treated group. Furthermore, TXA use was associated with a significantly lower risk of myocardial infarction (0.4% vs. 0.6%) and central nervous system failure (26.9% vs. 38.7%)(27).

Clinical guidelines, such as those of the National Institute for Health and Care Excellence (NICE), recommend its use in patients with severe traumatic hemorrhage. Thus, the use of TXA in the prehospital management of trauma patients indicates that its early administration, within the first hour, can reduce mortality. Administering it after three hours can increase the risk of adverse events, such as disseminated intravascular coagulation. Studies show variations in its effectiveness depending on the type of trauma and the time of intervention, as well as the impact on long-term functionality, so extensive and in-depth research related to the administration of TXA in the prehospital context is still needed (28,29,30).

## CONCLUSIONS

Prehospital TXA use in trauma patients has been shown to be effective in reducing short-term mortality, with significant results in the first 24 hours and at 28 days post-trauma. However, no significant improvements were seen in 6-month survival or long-term functional outcomes. Furthermore, no significant increase in the risk of serious adverse events such as deep vein thrombosis, pulmonary embolism, myocardial infarction, or ischemic stroke was identified compared with placebo.

## Data Availability

All data produced in the present work are contained in the manuscript

---

**Graphic 1.**
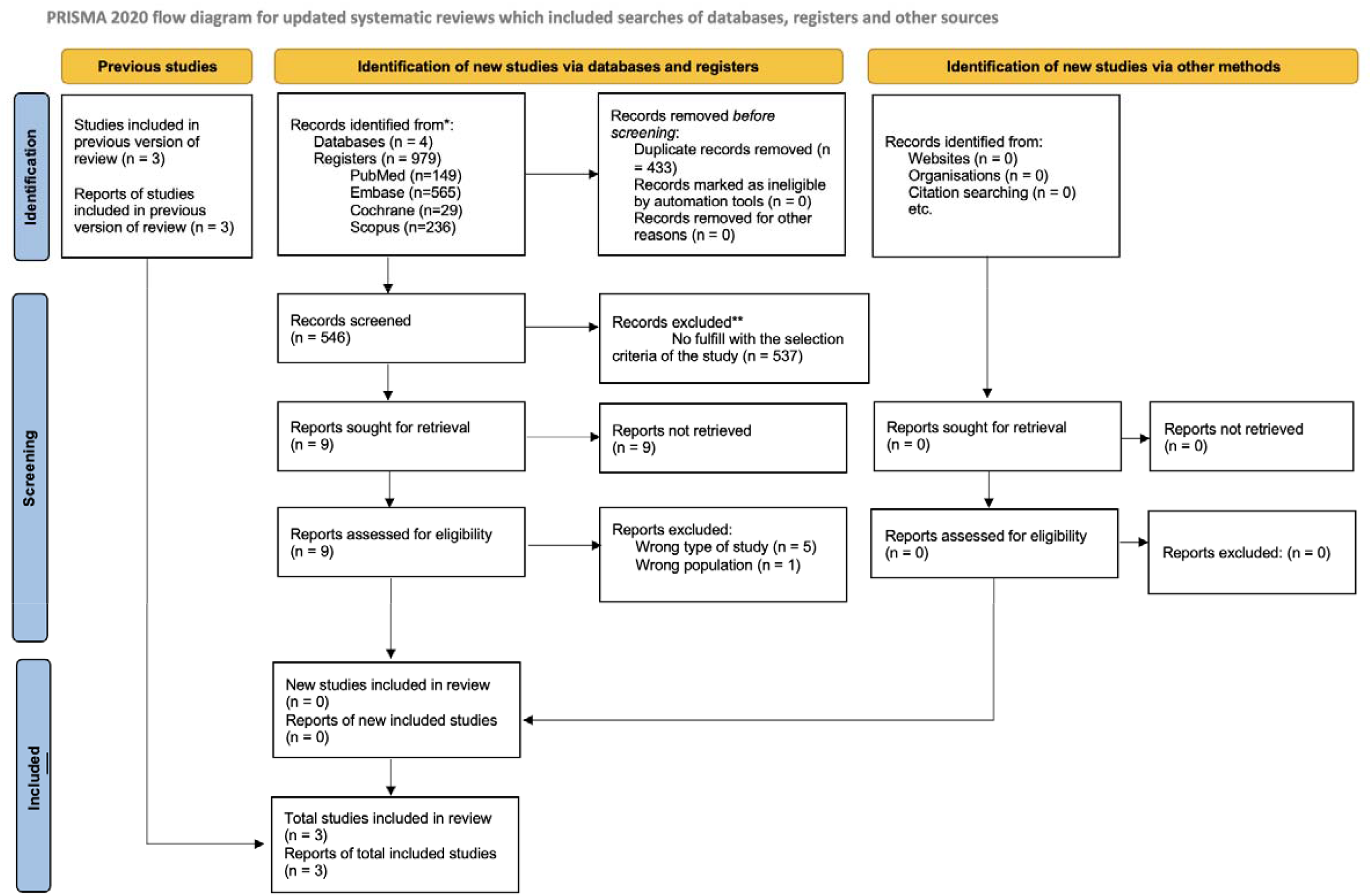
PRISMA Flow-diagram about the studies included in the review.

## Annex 1: Search strategy

### 1. Keywords

| Variable | Keywords |
| --- | --- |
| Prehospital | Emergency Medical Services<br>Out-of-Hospital Care<br>Prehospital Care |
| Tranexamic Acid | TXA<br>Antifibrinolytic patients |

### 2. Search Strategy

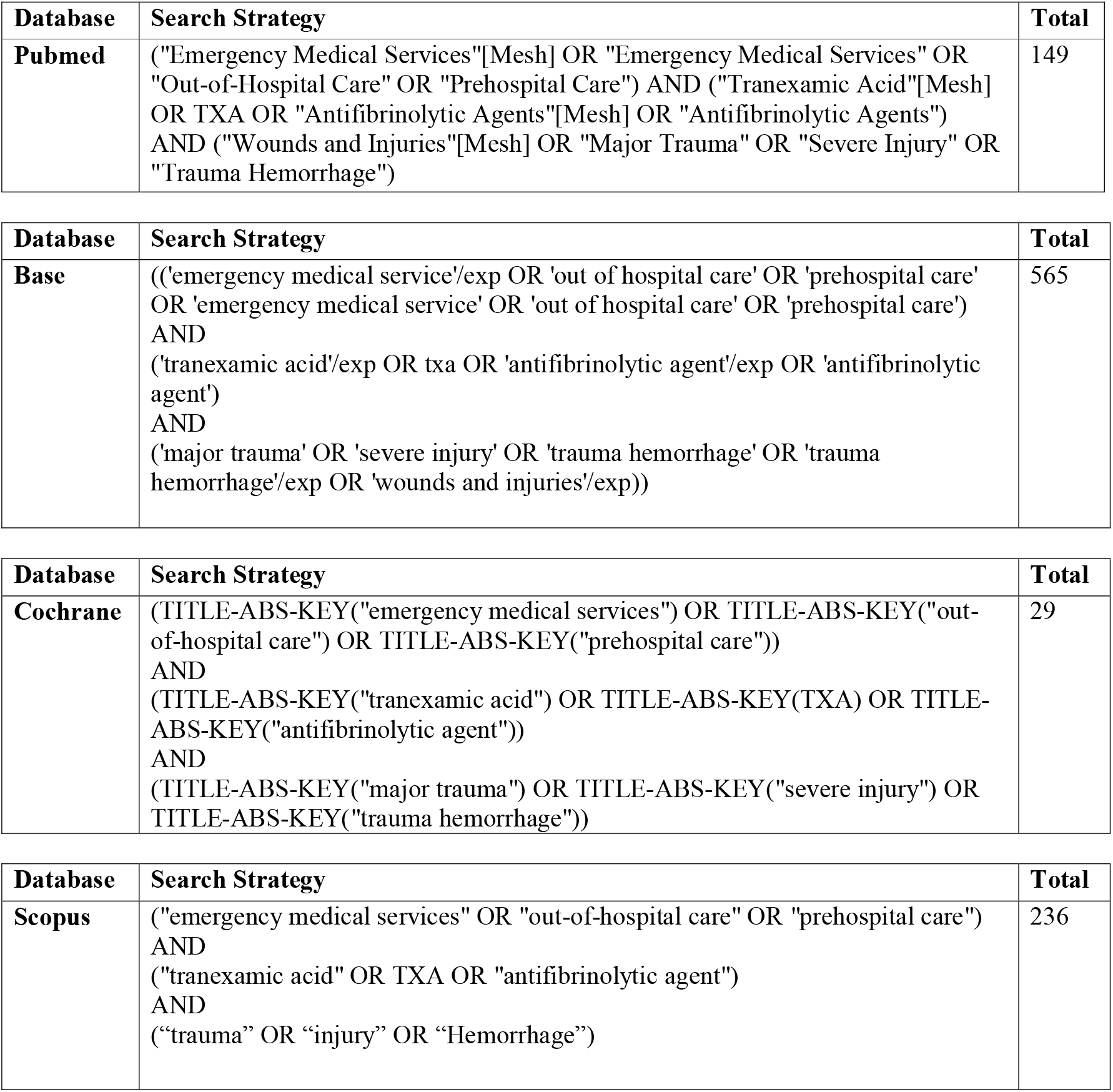

